# Data Resource Profile: Health Insurance Review and Assessment Service Korean Nationwide Claims OMOP-CDM (2015–2024) database, *“HIRA K-OMOP”*

**DOI:** 10.64898/2025.12.08.25341603

**Authors:** Dong Han Yu, Sun Mi Lim, Hyun Chul Shin, Moosung Kim, Seng Chan You, Rae Woong Park, Chungsoo Kim

## Abstract

The Health Insurance Review and Assessment Service Korean Nationwide Claims OMOP-CDM database (HIRA K-OMOP) is a nationwide data resource formatted according to the Observational Medical Outcomes Partnership (OMOP) Common Data Model (CDM) and derived from South Korea’s National Health Insurance claims data. It includes patient-level information and insurance claims for the entire population of South Korea from 2015 to 2024, providing population-scale coverage (56,416,773 patients). The HIRA K-OMOP database captures comprehensive data generated under the National Health Insurance program, including diagnoses, medications (inpatient and outpatient prescriptions and pharmacy dispensing records), procedures, and claim costs. This enables high-resolution observation of healthcare utilization and clinical practice among the Korean population.

The database was constructed by mapping Korean Electronic Data Interchange (EDI) codes to OMOP standard concepts. The requisite mapping tables were developed by the investigators of the “Integrating Local Vocabulary into OMOP CDM” study and released in June 2025, allowing nationally generated claims data to be transformed into an analysis-ready research resource. The complete analytic vocabulary set underpinning the build was obtained from the official OHDSI ATHENA portal (athena.ohdsi.org) in March 2025. Key strengths of the database include deterministic linkage to the national death registry and the explicit separation of prescribing and dispensing records, which enhances the validity of drug exposure assessments. The database is accessible to researchers through a privacy-preserving distributed research environment managed by HIRA.

**Key Features:**

- The Health Insurance Review and Assessment Service Korean Nationwide Claims OMOP-CDM database (*HIRA K-OMOP*) is a nationwide data resource formatted according to the Observational Medical Outcomes Partnership (OMOP) Common Data Model (CDM) and derived from South Korea’s national health insurance claims data.^1,2^
- It includes patient-level information and insurance claims for the entire population of South Korea from 2015 to 2024, providing population-scale coverage (56,416,773 patients).
- The *HIRA K-OMOP* database includes comprehensive data recorded as part of the National Health Insurance program, encompassing diagnosis, medications (inpatient, outpatient prescriptions, and dispensing records), procedure and claim costs. This allows for high-resolution observation of healthcare utilization and clinical practice among the Korean population.^3,4^
- It was constructed by mapping Korean Electronic Data Interchange (EDI) codes to OMOP standard concepts (concept IDs). The requisite mapping tables were developed by the investigators of the “Integrating Local Vocabulary into OMOP CDM” study and released in June 2025.^5^ Consequently, nationally generated claims data have been transformed into an analysis-ready research resource. The complete analytic vocabulary set, which underpins the build, was obtained from the official OHDSI ATHENA portal (athena.ohdsi.org) in March 2025.^6^

## DATA RESOURCE BASICS

### Role of HIRA in health system of South Korea

The health insurance system in the Republic of Korea is a mandatory social health insurance programme known as the National Health Insurance (NHI), which covers approximately 97% of citizens and residents.^7^ The NHI reimburses most medical expenses incurred during patient care, including diagnostic tests, medical procedures, and prescribed medications. The Health Insurance Review and Assessment Service (HIRA) is a quasi-governmental agency that plays a central role in this system by reviewing the appropriateness of services, evaluating quality of care, and monitoring healthcare utilisation and costs to ensure that care remains both effective and cost-efficient.

### HIRA K-OMOP Database

HIRA has access to a vast number of medical claims, making it an essential source of healthcare data in Korea. To promote data-driven research, HIRA has implemented an open data policy that allows claims data to be used for public purposes such as healthcare improvement and scientific research. Most requests are pre-processed by HIRA staff according to approved research protocols and then made available within a secure analytic environment, where researchers can access and analyses the data safely.

In 2023, HIRA developed a nationally representative COVID-19 database (*HIRA COVID-19 OMOP* database) covering approximately 10 million individuals and published its profile.^8^ The Observational Medical Outcomes Partnership (OMOP) Common Data Model (CDM), maintained by the Observational Health Data Sciences and Informatics (OHDSI), enables international distributed data analysis by applying the same query to healthcare data from multiple institutions. Recently, HIRA expanded the OMOP architecture to a population-wide scale, thereby supporting national-level analyses and aligning the database with emerging healthcare research priorities.^9,10^

### Separation of prescribing and dispensing in Korea

Since July 2000, South Korea has strictly enforced a system that separates prescribing from dispensing. Physicians are responsible for providing medical care and issuing prescriptions, while pharmacists are responsible for supplying and dispensing medications. Under this system, patients may choose not to have a prescription filled, and pharmacists are allowed to substitute prescribed medication with bioequivalent products. As a result, out-of-hospital prescription records issued by healthcare providers and dispensing records from local pharmacies are largely overlapping but are not fully identical. Because all pharmacies submit insurance claims for medication dispensing to HIRA, the agency can accurately capture nationwide dispensing data. The *HIRA K-OMOP* database is designed to provide both provider-level prescribing information and patient-centric medication dispensing records. To prevent duplication in analyses, out-of-hospital prescription records and dispensing records are maintained in separate schemas (Figure 1).

**Figure 1.**
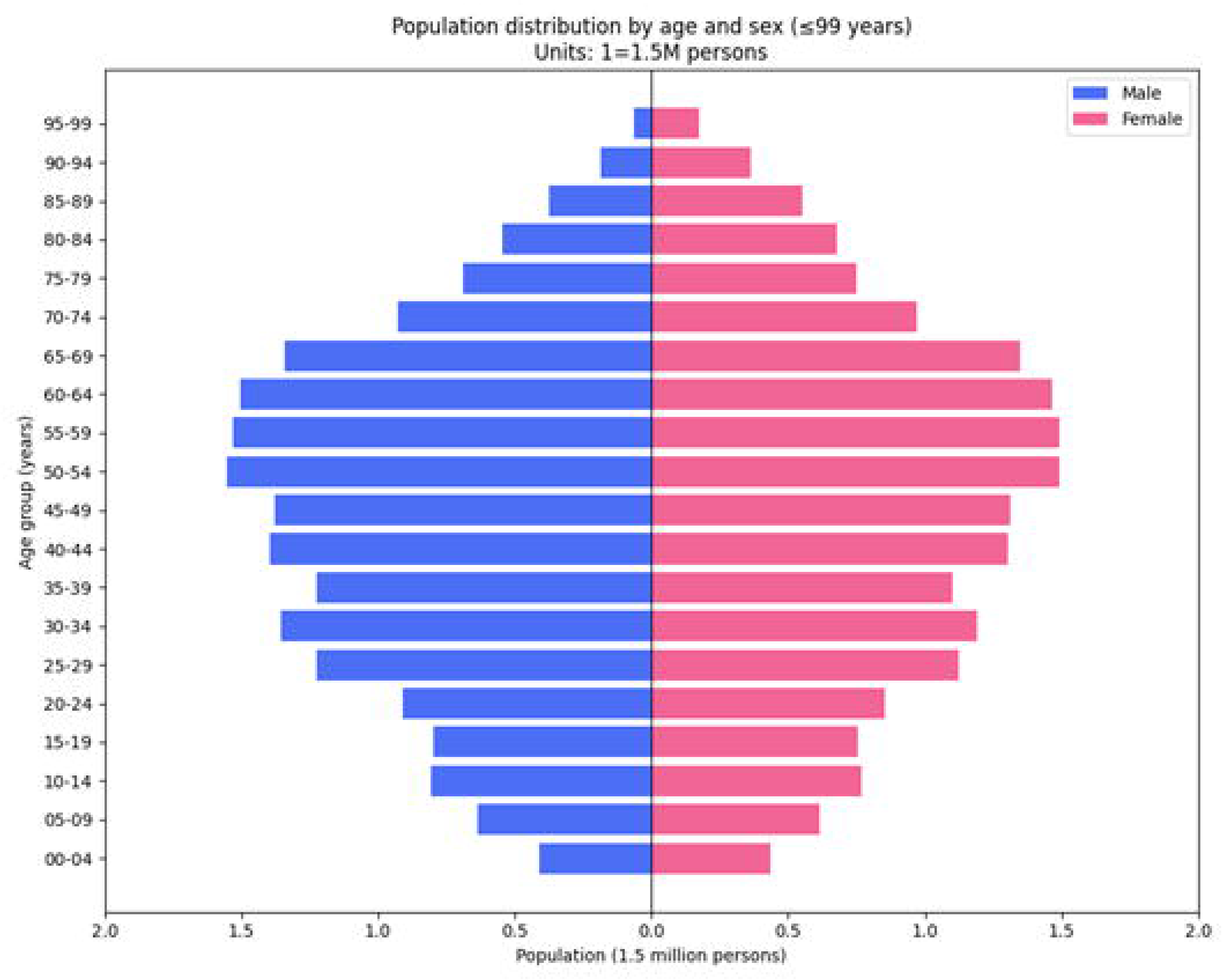
Composition of source data for building *HIRA K-OMOP* database & Clinical domain composition of *HIRA K-OMOP* database.

### Population coverage

The *HIRA K-OMOP* database was constructed using claims data for the entire NHI-covered population. It contains comprehensive medical records for 56 million insured patients.

### Frequency of data collection

Claims data are collected continuously as part of routine NHI operations. The *HIRA K-OMOP* database is refreshed on a biennial basis, with additional updates performed as needed for specific research initiatives.

### Privacy policy

The *HIRA K-OMOP* database has advantages in terms of personal information protection. The identification numbers of insurance subscribers managed by HIRA are replaced with serial numbers with matching digits when transformed into OMOP–CDM. Each subscriber’s original identification number is replaced with a surrogate key generated via a one-way random sequence assignment, in which unique random numbers are allocated and the lookup table is subsequently destroyed. As a result, there is no way to reconstruct the original IDs, and it is impossible to identify specific individuals from the transformed database.

## DATA COLLECTED

### Data source

A schematic diagram of the data source is presented in Figure 1. The dataset was obtained from the data warehouse operated by HIRA, which includes national insurance claims that were completed through the official review process between January 2015 and December 2024, spanning a total of ten years. The HIRA claims database is organized into several core tables including Provider Claim Information (200 table), Pharmacy Claim Information (201 table), Treatment Details (300 table), Disease Information (400 table), Out-of-Hospital Prescription Details (530 table), Pharmacy Dispensing Details (301 table), and Healthcare Institution Status Information.

The claim specification table contains key administrative and clinical attributes for each claim, such as the claim key, patient surrogate key, healthcare institution key, primary and secondary disease codes, treatment start and end dates, duration of treatment, insurer liability, and visit details. The Treatment Details table records procedures, medications, and treatment materials with their respective codes, dosage amounts, and administration frequencies. The disease table is defined at the claim-statement level and records up to five diagnoses per claim—one principal and up to four secondary—aligned to the relevant clinical department. The Out-of-Hospital Prescription Details table captures dosage, administration frequency, and treatment duration for medications prescribed outside hospitals. The Pharmacy Claim Information table records pharmacy-submitted claims and classifies them as either extemporaneous compounding or prescription-based dispensing. The Pharmacy Dispensing Details table records charges for pharmacist services (e.g., basic dispensing fees and medication counselling fees). It also includes product-level records for medicines prepared extemporaneously or dispensed against an issued prescription. The Healthcare Institution Status Information table provides information on institution identifiers, regional locations, and institution types. Mortality data were linked from the national death registry maintained by Statistics Korea.

### Extract, Transform, Load

The target population consisted of all patients whose insurance claims had been reviewed between 2015 and 2024. Based on this population, claims data from January 1, 2015, to December 31, 2024 were extracted. The extracted dataset included a wide range of key patient information, including demographics (sex, age, type of insurance), visit type (outpatient or inpatient care), medical history (diagnoses, procedures, and treatments), examination history, and prescription and dispensing records (such as the number of days prescribed and list of medications dispensed). The detailed specifications of these key variables are provided in Table 1.

**Table 1.**
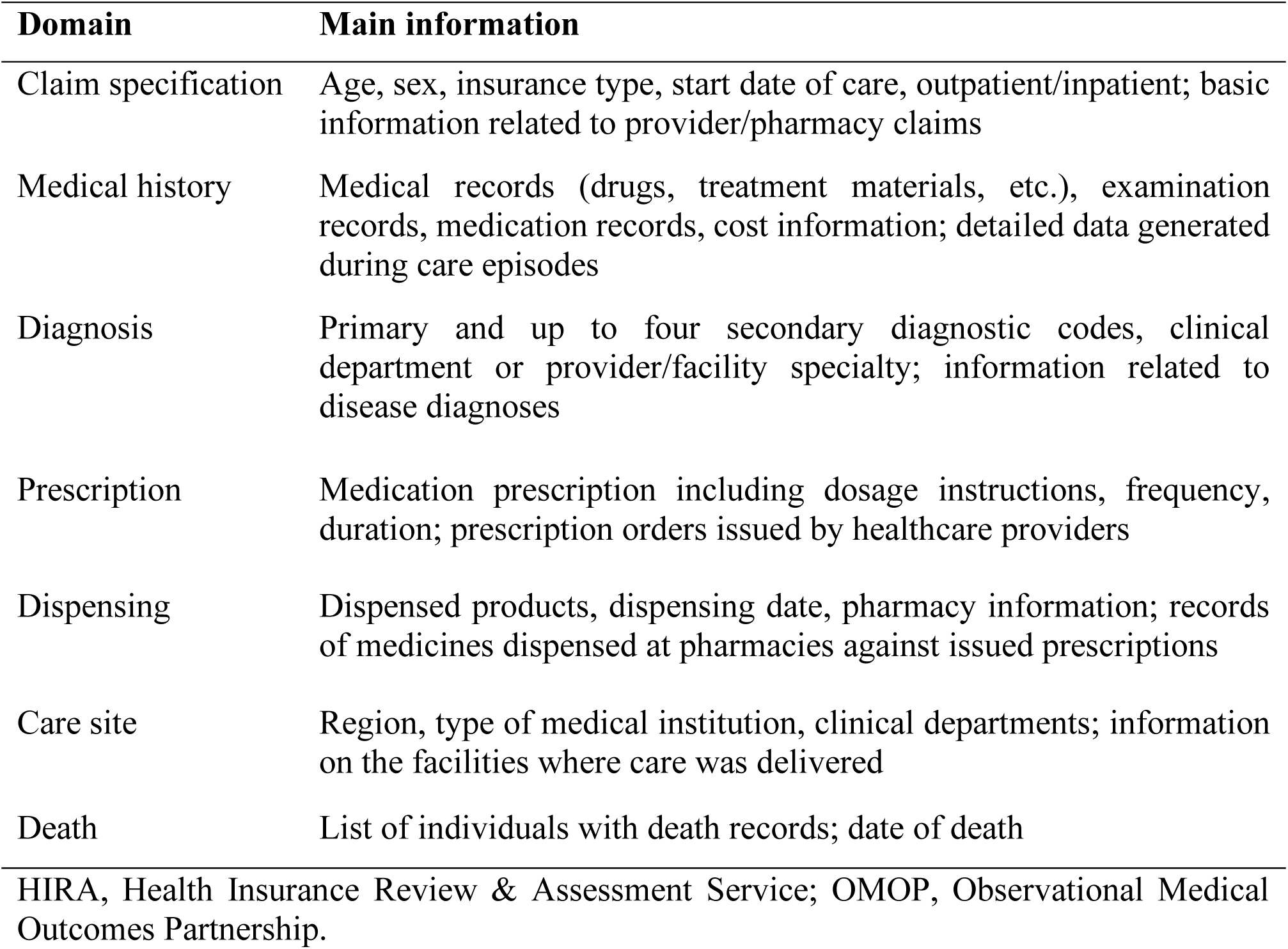
Key information included in the *HIRA K-OMOP* database.

| Domain | Main information |
| --- | --- |
| Claim specification | Age, sex, insurance type, start date of care, outpatient/inpatient; basic information related to provider/pharmacy claims |
| Medical history | Medical records (drugs, treatment materials, etc.), examination records, medication records, cost information; detailed data generated during care episodes |
| Diagnosis | Primary and up to four secondary diagnostic codes, clinical department or provider/facility specialty; information related to disease diagnoses |
| Prescription | Medication prescription including dosage instructions, frequency, duration; prescription orders issued by healthcare providers |
| Dispensing | Dispensed products, dispensing date, pharmacy information; records of medicines dispensed at pharmacies against issued prescriptions |
| Care site | Region, type of medical institution, clinical departments; information on the facilities where care was delivered |
| Death | List of individuals with death records; date of death |

HIRA, Health Insurance Review & Assessment Service; OMOP, Observational Medical Outcomes Partnership.

As shown in Figure 1, the extracted claims data were then transformed into the OMOP CDM version 5.3, enabling meaningful clinical information to be represented in a standardized structure suitable for large-scale observational research.^11^ This transformation involved the construction of several primary OMOP tables, such as PERSON, CONDITION_OCCURRENCE, DRUG_EXPOSURE and COST, which are widely used in real-world evidence studies.

For terminology standardization, each domain table utilized the concept IDs from the OMOP standardized vocabulary. The mapping of Korean claims codes to OMOP concepts was performed using the EDI-SNOMED and EDI-RxNorm vocabulary mapping dictionaries applied by HIRA in March 2025.

### Data distribution

Table 2 presents the number of individuals and records included in 17 data tables, excluding vocabulary tables. In total, the *HIRA K-OMOP* database includes information on 56,416,773 insured persons. The CONDITION_OCCURRENCE table, which stores all diagnosis records for these individuals, contains 28,150,894,981 entries. The DRUG_EXPOSURE table includes 26,966,367,283 records of prescribed medications, while the DRUG_EXPOSURE_D (Dispensing) table contains 19,553,296,933 dispensing events. The PROCEDURE_OCCURRENCE table has the largest volume, with 16,281,745,905 documented procedures.

**Table 2.**
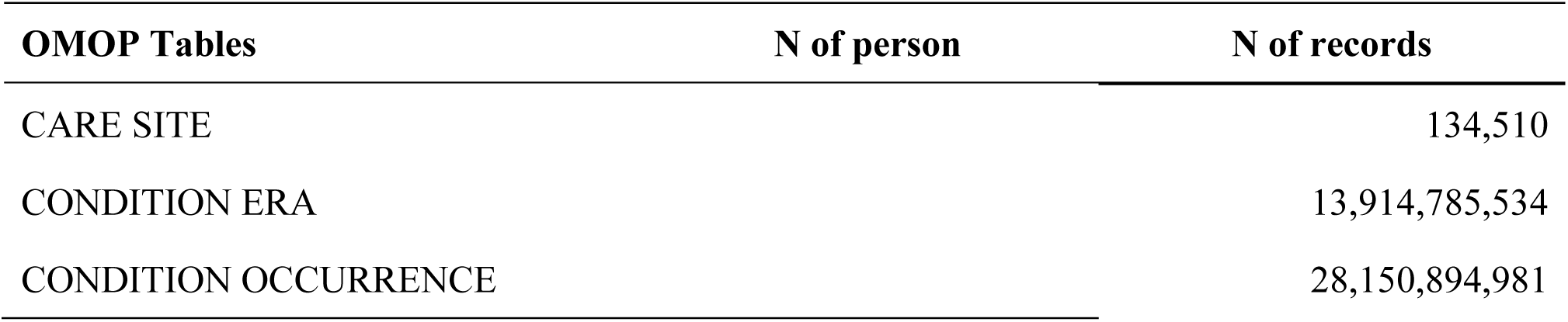
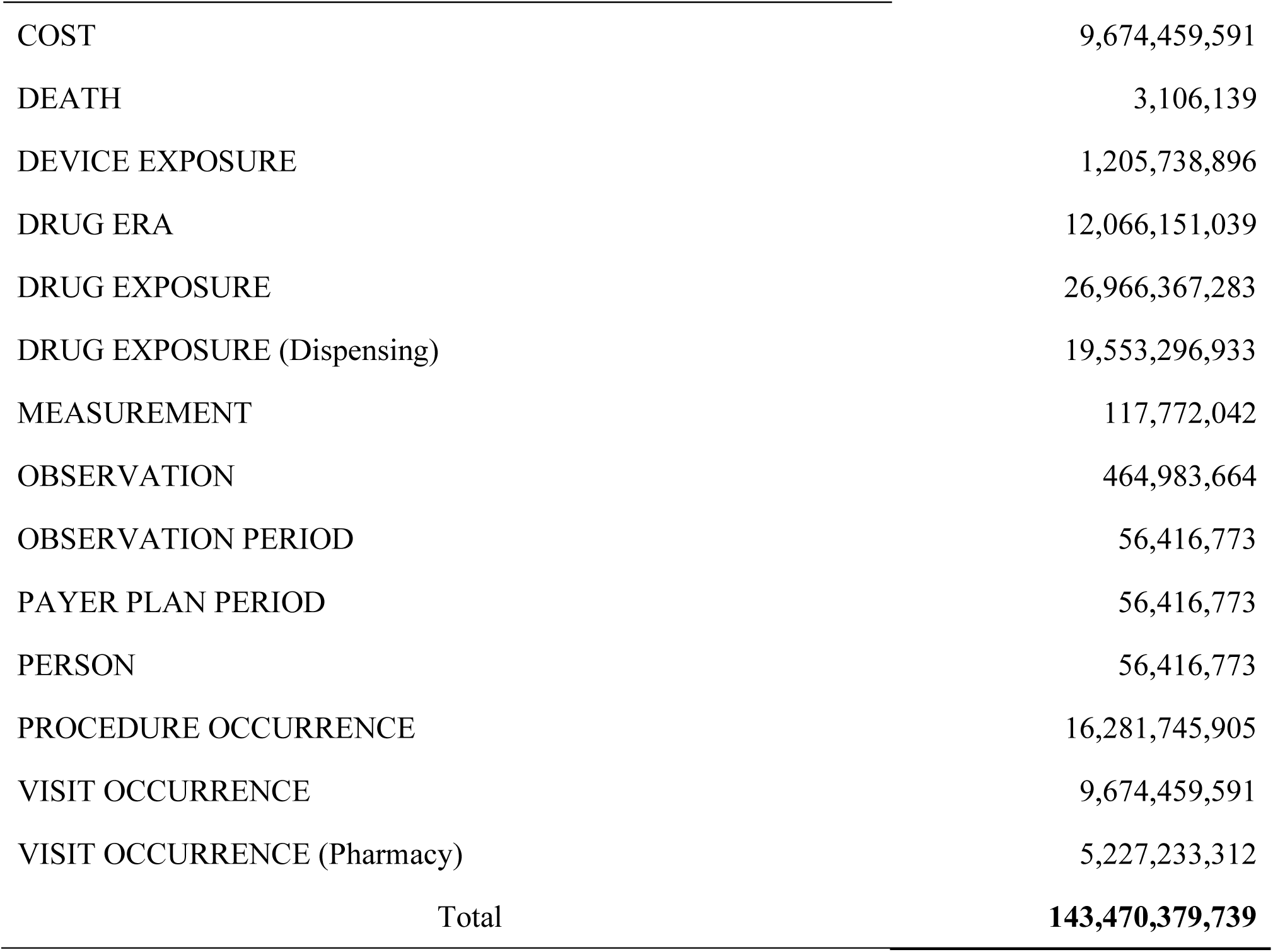
Number of persons and records in the *HIRA K-OMOP* database.

| OMOP Tables | N of person | N of records |
| --- | --- | --- |
| CARE SITE |  | 134,510 |
| CONDITION ERA |  | 13,914,785,534 |
| CONDITION OCCURRENCE |  | 28,150,894,981 |

|  |  |
| --- | --- |
| COST | 9,674,459,591 |
| DEATH | 3,106,139 |
| DEVICE EXPOSURE | 1,205,738,896 |
| DRUG ERA | 12,066,151,039 |
| DRUG EXPOSURE | 26,966,367,283 |
| DRUG EXPOSURE (Dispensing) | 19,553,296,933 |
| MEASUREMENT | 117,772,042 |
| OBSERVATION | 464,983,664 |
| OBSERVATION PERIOD | 56,416,773 |
| PAYER PLAN PERIOD | 56,416,773 |
| PERSON | 56,416,773 |
| PROCEDURE OCCURRENCE | 16,281,745,905 |
| VISIT OCCURRENCE | 9,674,459,591 |
| VISIT OCCURRENCE (Pharmacy) | 5,227,233,312 |
| Total | **143,470,379,739** |

The baseline characteristics of the overall population are summarized in Table S1. Among all insured individuals, 49.86% were women. Figure 2 displays the age-sex distribution of the population, grouped in five-year age intervals. The overall mortality proportion recorded in the database was 5.51%.

**Figure 2.**
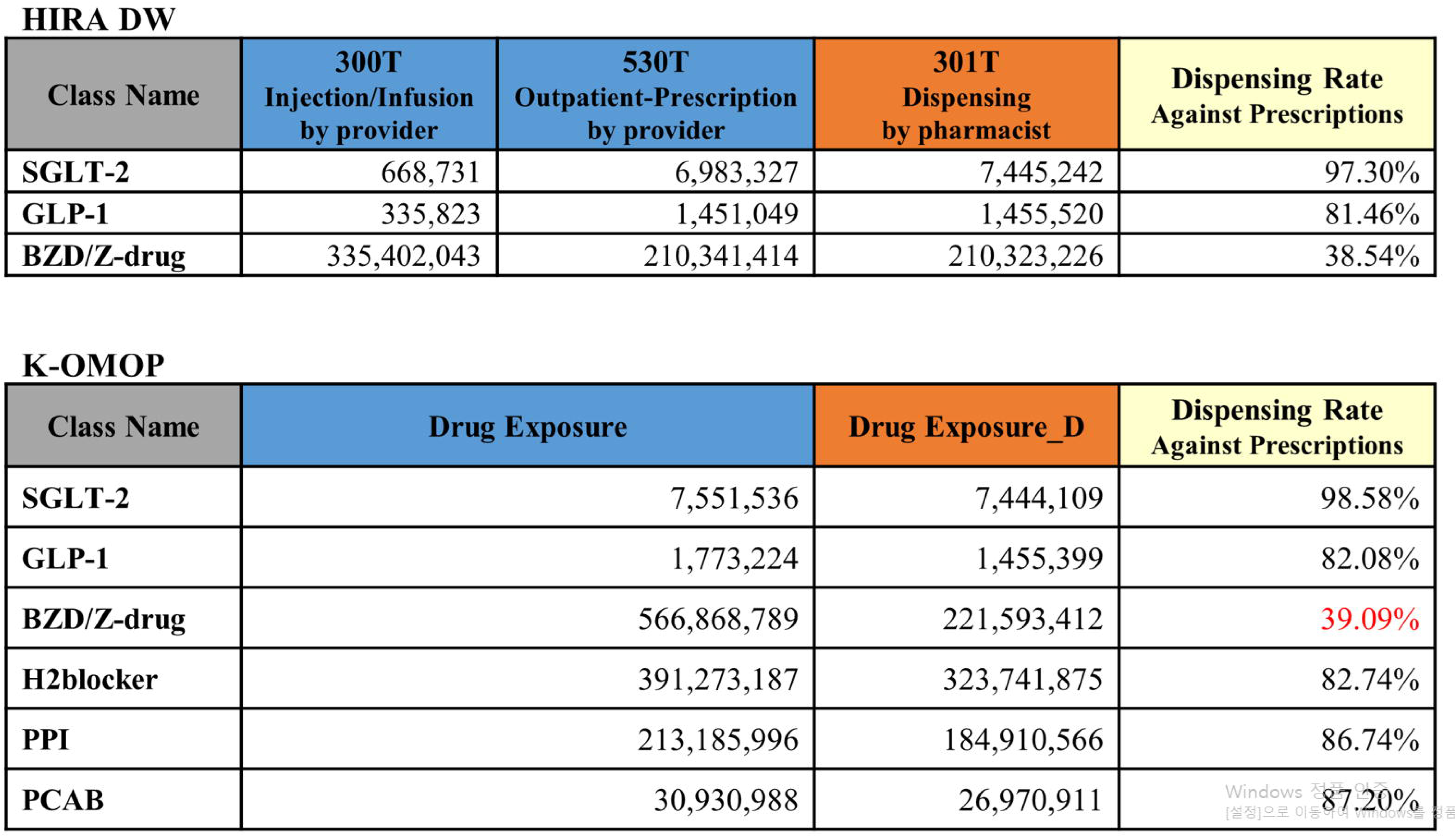
Population distribution by sex and 5-year-old age group.

### Data Resource Use

HIRA has successfully completed two pilot programs to provide controlled access to OMOP CDM data. The first pilot, conducted from July 2022 to June 2023, utilized the *COVID-19 OMOP* database and supported more than 40 research projects under a distributed research model. The second pilot, carried out from September 2023 to December 2024, implemented the newly developed *HIRA K-OMOP* database and facilitated over 15 projects. The research outputs from these pilots included studies examining the associations between COVID-19 and specific health conditions, as well as nationwide evaluations of drug effectiveness and safety.^12,13^

### Strengths and weaknesses

i. Population-scale coverage with outcome ascertainment and international interoperability. The *HIRA K-OMOP* database encompasses virtually the entire population of South Korea across outpatient and inpatient settings, enabling large-scale pharmacoepidemiology and health-services research. Deterministic linkage to the national death registry facilitates robust mortality outcome ascertainment. Conversion to the OMOP Common Data Model (CDM) enhances reproducibility and cross-database comparability and thus supports multi-institutional and multinational research.^14–17^
ii. Explicit separation of prescribing and dispensing to reduce drug-exposure misclassification. Korea’s separation of prescribing and dispensing is operationalised by recording provider prescribing and pharmacy dispensing as distinct events, thereby mitigating double counting and improving the validity of drug-exposure definitions. Figure 3 compares dispensing-to-prescribing ratios across six clinically relevant drug classes. Dispensing-to-prescribing ratios were approximately 98% for SGLT2 inhibitors, around 82% for GLP-1 receptor agonists (with roughly 18 % of issued prescriptions not filled), and only about 39 % for benzodiazepines and Z-drugs. These patterns underscore the value of dispensing data within *HIRA K-OMOP* for capturing actual medication use and reducing exposure misclassification.
iii. Privacy-preserving, governance-ready research operations. The database is accessed within a secure, distributed research environment. Direct identifiers are never exposed, and export of patient-, claim- or institution-level granular statistics is restricted. Together with IRB and HIRA approvals, these controls provide a governance framework that balances analytical utility with rigorous protection of personal information.^18^

However, the current data resource also has several limitations. To more fully reflect real-world clinical practice, additional data would be valuable, including non-reimbursed services, over-the-counter (OTC) medications, vaccines, and laboratory test results. Because HIRA cannot capture items that are not reimbursed by the NHI or OTC drugs sold directly at pharmacies without prescriptions, important clinical exposures may be missing. This gap has the potential to act as a major confounder in healthcare research. For example, the use of OTC medications such as acetaminophen or non-steroidal anti-inflammatory drugs (NSAIDs) cannot be accounted for when assessing drug-related risks. In addition, due to the inherent nature of the claims data, the *HIRA K-OMOP* database does not include laboratory results, which limits the ability to conduct studies that require laboratory values, such as biomarker-based analyses or research involving disease severity and treatment response.^19,20^

**Figure 3.**
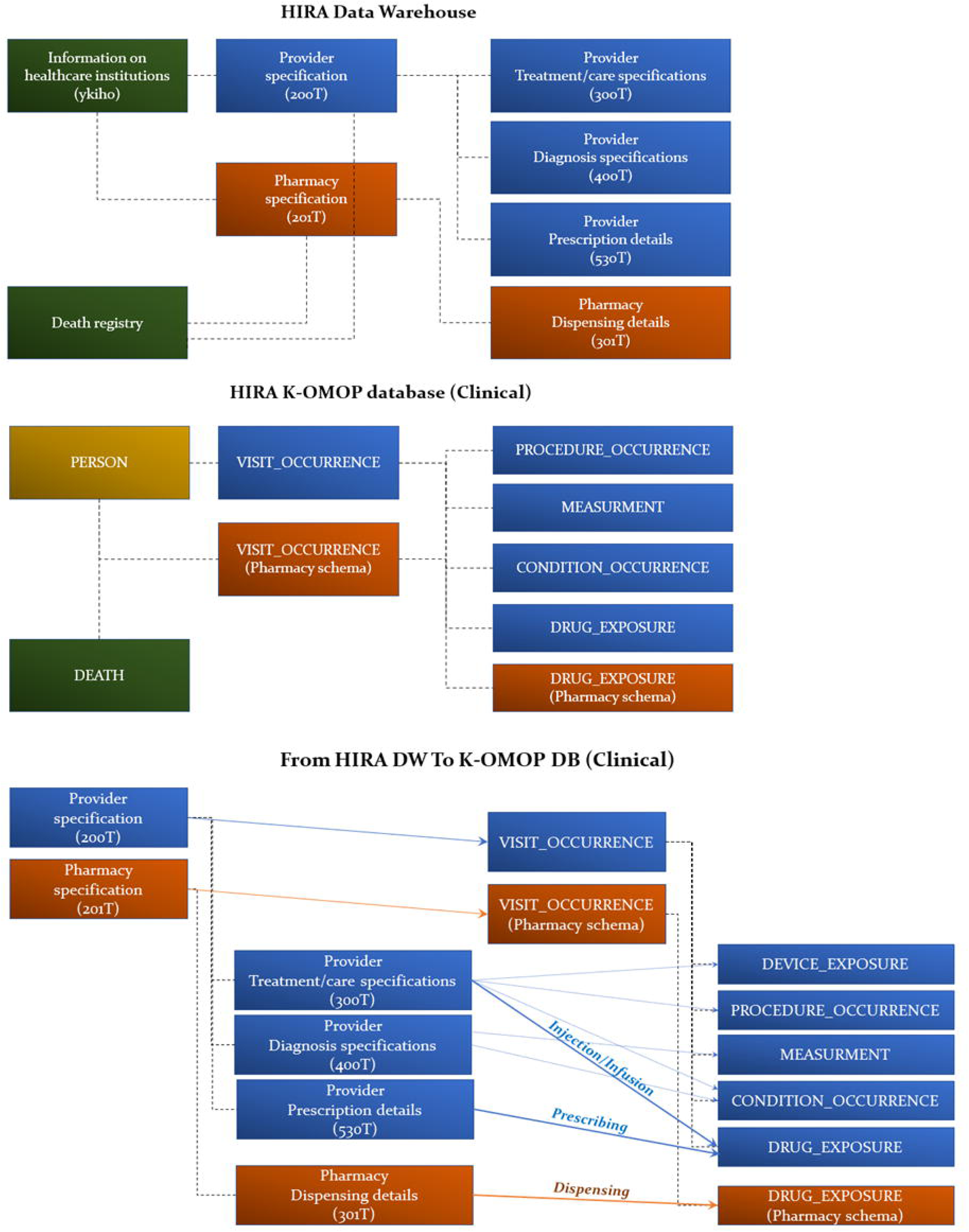
Comparison of Prescribing and Dispensing Outcomes for Three Specific Drug Classes.

Nevertheless, such limitations can be effectively mitigated by utilizing the *HIRA K-OMOP* database in conjunction with hospital-based CDMs, such as the Federated E-health Big Data for Evidence Renovation Network (FEEDER-NET).^21^ While FEEDER-NET comprises hospital Electronic Medical Records (EMR) containing detailed clinical data—including laboratory results and non-reimbursed services—it operates as a distributed network where patient linkage across institutions is not feasible. Therefore, a comparative analysis that leverages both the population-wide coverage of *HIRA K-OMOP* and the deep clinical granularity of hospital CDMs would offer a robust strategy for generating real-world evidence. Indeed, the analytic value of HIRA CDM has already been validated through its use in studies published in major journals, further underscoring its potential for high-impact research.^22,23^

### Data Resource Access

The database is maintained by the Big Data Department of the Health Insurance Review and Assessment Service (HIRA), and inquiries regarding its use should be directed to. Access for domestic researchers requires approval from an institutional review board as well as authorisation from HIRA.

## Data Availability

The HIRA K-OMOP database analysed in this study is not publicly available because of legal and privacy restrictions on Korean National Health Insurance claims data. Researchers may request access through the Health Insurance Review and Assessment Service (HIRA) data provision service on a project-by-project basis, after obtaining approval from an appropriate institutional review board and HIRA's data access committee. Analyses are conducted within HIRA's secure research environment, and only aggregate, de-identified results that pass disclosure control can be exported.

## Ethics approval

The HIRA is a quasi-governmental organization authorized under the relevant laws of the Republic of Korea to process personal information. Since the construction of the database itself did not constitute human subjects research, it was exempt from review by an institutional bioethics committee. However, for any distributed research conducted by individual investigators, submission to and approval from an institutional review board is mandatory.

## Author contributions

D H Yu designed the conversion logic of the database, performed the data conversion, and wrote the manuscript. S M Lim and H C Shin validated the converted data using the Data Quality Dashboard (DQD).^24,25^ M Kim reviewed data security for the converted database. S C You and R W Park provided the vocabulary mapping sets. C S Kim supervised all processes. All authors reviewed the manuscript and provided critical revisions.

## Acknowledgements

This work was supported by the Health Insurance Review and Assessment Service (HIRA). The views expressed are those of the authors and not necessarily those of the HIRA.

## References

1. Overhage JM, Ryan PB, Reich CG, Hartzema AG, Stang PE. Validation of a common data model for active safety surveillance research. J Am Med Inform Assoc JAMIA. 2012;19(1):54–60.

2. Reinecke I, Zoch M, Reich C, Sedlmayr M, Bathelt F. The Usage of OHDSI OMOP - A Scoping Review. Stud Health Technol Inform. 2021 Sept 21;283:95–103.

3. Yoon D, Ahn EK, Park MY, et al. Conversion and Data Quality Assessment of Electronic Health Record Data at a Korean Tertiary Teaching Hospital to a Common Data Model for Distributed Network Research. Healthc Inform Res. 2016 Jan;22(1):54–58.

4. Hripcsak G, Ryan PB, Duke JD, et al. Characterizing treatment pathways at scale using the OHDSI network. Proc Natl Acad Sci U S A. 2016 July 5;113(27):7329–7336.

5. Park Y, Yoon J, Zhuk A, Ostropolets A, You SC. Integrating Local Vocabulary into OMOP CDM: A Step-by-Step Tutorial [Internet]. medRxiv; 2025 [cited 2025 Nov 21]. p. 2025.05.07.25327200. Available from: https://www.medrxiv.org/content/10.1101/2025.05.07.25327200v1

6. Reich C, Ostropolets A, Ryan P, et al. OHDSI Standardized Vocabularies-a large-scale centralized reference ontology for international data harmonization. J Am Med Inform Assoc JAMIA. 2024 Feb 16;31(3):583–590.

7. Cheol Seong S, Kim Y-Y, Khang Y-H, et al. Data Resource Profile: The National Health Information Database of the National Health Insurance Service in South Korea. Int J Epidemiol. 2017 June 1;46(3):799–800.

8. Kim C, Yu DH, Baek H, Cho J, You SC, Park RW. Data Resource Profile: Health Insurance Review and Assessment Service Covid-19 Observational Medical Outcomes Partnership (HIRA Covid-19 OMOP) database in South Korea. Int J Epidemiol. 2024 Apr 11;53(3):dyae062.

9. Hripcsak G, Duke JD, Shah NH, et al. Observational Health Data Sciences and Informatics (OHDSI): Opportunities for Observational Researchers. Stud Health Technol Inform. 2015;216:574–578.

10. Haendel MA, Chute CG, Bennett TD, et al. The National COVID Cohort Collaborative (N3C): Rationale, design, infrastructure, and deployment. J Am Med Inform Assoc JAMIA. 2021 Mar 1;28(3):427–443.

11. Makadia R, Ryan PB. Transforming the Premier Perspective® Hospital Database into the Observational Medical Outcomes Partnership (OMOP) Common Data Model. eGEMs. 2014 Nov 11;2(1):1110.

12. Matcho A, Ryan P, Fife D, Reich C. Fidelity assessment of a clinical practice research datalink conversion to the OMOP common data model. Drug Saf. 2014 Nov;37(11):945–959.

13. Ryan PB, Schuemie MJ, Welebob E, Duke J, Valentine S, Hartzema AG. Defining a reference set to support methodological research in drug safety. Drug Saf. 2013 Oct;36 **Suppl 1**:S33–47.

14. Voss EA, Makadia R, Matcho A, et al. Feasibility and utility of applications of the common data model to multiple, disparate observational health databases. J Am Med Inform Assoc JAMIA. 2015 May;22(3):553–564.

15. Schuemie MJ, Gini R, Coloma PM, et al. Replication of the OMOP experiment in Europe: evaluating methods for risk identification in electronic health record databases. Drug Saf. 2013 Oct;36 **Suppl 1**:S159–169.

16. Kostka K, Duarte-Salles T, Prats-Uribe A, et al. Unraveling COVID-19: A Large-Scale Characterization of 4.5 Million COVID-19 Cases Using CHARYBDIS. Clin Epidemiol. 2022;14:369–384.

17. Principles of Large-scale Evidence Generation and Evaluation across a Network of Databases (LEGEND) | Journal of the American Medical Informatics Association | Oxford Academic [Internet]. [cited 2025 Nov 21]. Available from: https://academic.oup.com/jamia/article/27/8/1331/5895561

18. Lane JCE, Weaver J, Kostka K, et al. Risk of hydroxychloroquine alone and in combination with azithromycin in the treatment of rheumatoid arthritis: a multinational, retrospective study. Lancet Rheumatol. 2020 Nov;2(11):e698–e711.

19. Biedermann P, Ong R, Davydov A, et al. Standardizing registry data to the OMOP Common Data Model: experience from three pulmonary hypertension databases. BMC Med Res Methodol. 2021 Nov 2;21(1):238.

20. Converting to a Common Data Model: What is Lost in Translation? | Drug Safety [Internet]. [cited 2025 Nov 21]. Available from: https://link.springer.com/article/10.1007/s40264-014-0221-4

21. You SC, Lee S, Choi B, Park RW. Establishment of an International Evidence Sharing Network Through Common Data Model for Cardiovascular Research. Korean Circ J. 2022 Dec;52(12):853–864.

22. Burn E, You SC, Sena AG, et al. Deep phenotyping of 34,128 adult patients hospitalised with COVID-19 in an international network study. Nat Commun. Nature Publishing Group; 2020 Oct 6;11(1):5009.

23. Morales DR, Conover MM, You SC, et al. Renin–angiotensin system blockers and susceptibility to COVID-19: an international, open science, cohort analysis. Lancet Digit Health. Elsevier; 2021 Feb 1;3(2):e98–e114.

24. Blacketer C, Defalco FJ, Ryan PB, Rijnbeek PR. Increasing trust in real-world evidence through evaluation of observational data quality. J Am Med Inform Assoc JAMIA. 2021 Sept 18;28(10):2251–2257.

25. Kahn MG, Callahan TJ, Barnard J, et al. A Harmonized Data Quality Assessment Terminology and Framework for the Secondary Use of Electronic Health Record Data. eGEMs. 2016 Sept 11;4(1):1244.

