## Supplementary material for "Data Resource Profile: Health Insurance Review and Assessment Service Korean Nationwide Claims OMOP-CDM (2015–2024) database, *“HIRA K-OMOP”*"

**Table S1. Basic characteristics of *HIRA K-OMOP* database**

| | **Total** |
|---|---|
| N | **56,416,773** |
| Sex, n (%) | |
| Female | 28,127,895 (49.9) |
| Male | 28,288,878 (50.1) |
| N of patients at the midyear | |
| 2015 | 48,492,906 |
| 2016 | 48,991,244 |
| 2017 | 49,160,835 |
| 2018 | 49,460,680 |
| 2019 | 49,623,312 |
| 2020 | 48,561,259 |
| 2021 | 49,150,210 |
| 2022 | 50,610,003 |
| 2023 | 50,297,359 |
| 2024 | 50,039,596 |
| Age group*, n (%) | |
| 0-19 | 7,839,663 (13.9) |
| 20-39 | 13,472,509 (23.9) |
| 40-59 | 17,181,427 (30.5) |
| 60-79 | 13,492,474 (23.9) |
| ≥ 80 | 4,430,700 (7.9) |
| Visit, n (%) | |
| Inpatient | 163,280,924 (1.7) |
| Outpatient | 9,482,046,621 (98.0) |
| Emergency Room | 344,130 (0.003) |
| Emergency Room and Inpatient | 28,787,916 (0.3) |
| Visit_P, n (%) | |
| Pharmacy visit | 5,227,233,312 (100) |
| Death, n (%) | 3,106,139 (5.5) |

HIRA: health insurance review and assessment service; *Age group is defined based on the age at midyear of 2025.

### Table S2. Contents of data tables in *HIRA K-OMOP* database

| Data tables | Contents / **Fields** |
|---|---|
| PERSON | One record per unique beneficiary enrolled in the National Health Insurance (NHI). Represents the analytic "patient" population of HIRA K-OMOP from 2015–2024. / **Sex, Date of birth (year, month, day) etc.** |
| OBSERVATION_PERIOD | Time intervals during which a person is under continuous NHI coverage and thus has complete capture of reimbursed healthcare utilization in HIRA claims. / O**bservation period start and end dates, period type etc.** |
| DEATH | The table contains the clinical event for how and when a Person dies. A person can have up to one record if the source system contains evidence about the Death. / **Date of death, Death type (Death Certificate: 32507) etc.** |
| VISIT_OCCURRENCE | Claim-level visits derived from individual claim records in the source data. Each record represents a billed encounter at a healthcare institution (hospital, clinic, public health center, etc.) and is linked to diagnoses, procedures, and prescriptions. / **Visit concept (Inpatient: 9201, Outpatient: 9202, Emergency department visit: 9203, Emergency department and Inpatient visit: 262), visit start date, end date, visit type (Claim: 32810), care site etc.** |
| VISIT_OCCURRENCE_P | Claim-level visits derived from individual claim records in the source data. Each record represents a billed encounter at a healthcare institution (pharmacy) and is linked to diagnoses, procedures, and prescriptions. / **Visit concept (Pharmacy visit: 581458), visit start date, end date, visit type (Claim: 32810), care site etc.** |
| PROCEDURE_OCCURRENCE | Procedures, surgeries, and other billable medical services performed during visits, extracted from procedure and treatment claim lines and mapped from Korean EDI procedure codes to standard concepts. **/ Procedure concept, procedure date etc.** |
| DRUG_EXPOSURE | Provider-level prescribing records captured from medical service claims. Each record reflects a medication ordered by a healthcare provider, irrespective of whether it was subsequently dispensed at a pharmacy. Source EDI drug codes are mapped to OMOP standard drug concepts (Rx-Norm). / **Drug concept, exposure start date, end date, quantity, days supply, total paid(non-standard) etc.** |
| DRUG_EXPOSURE_D | Pharmacy dispensing records captured from pharmacy claims. Represents drug exposures based on medications that were actually dispensed at community or hospital pharmacies, enabling comparison between prescribed and dispensed therapies and reducing exposure misclassification. / **Drug concept, exposure start date, end date, quantity, days supply, total paid(non-standard) etc.** |
| DEVICE_EXPOSURE | Billable medical devices and supplies (e.g. implantable devices, durable medical equipment, consumables) recorded on claims and mapped to standard device concepts where available. / **Device concept, exposure start date, end date, quantity, total paid(non-standard) etc.** |
| CONDITION_OCCURRENCE | All diagnosis codes reported on reimbursement claims, including principal and secondary diagnoses recorded during each visit episode. Source Korean EDI diagnosis codes are mapped to SNOMED CT standard concepts. / **Condition concept, condition start date, end date, condition type concept (primary: 44786627, 2nd: 44786629, 3rd: 45756845, 4th: 45756846, 5th: 45756847)** |
| MEASUREMENT | Diagnostic tests and laboratory examinations ordered and billed in claims (e.g. laboratory tests, imaging procedures) recorded as measurement events. Numeric result |

| | |
|---|---|
| | values are not available in HIRA claims, so the table primarily captures that the test was performed and when it occurred.<br>/ **Measurement concept, date, total paid(non-standard), value as number (NULL) etc.** |
| OBSERVATION | Other clinical or administrative facts not recorded as diagnoses or procedures, such as program eligibility flags, specific benefit classifications, or health screening indicators that can be mapped to observation concepts.<br>/ **Observation concept, date etc.** |
| CARE_SITE | Healthcare institutions where services were delivered, including tertiary hospitals, secondary hospitals, clinics, public health centers, and pharmacies.<br>/ **Care-site name, place of service concept etc.** |
| PAYER_PLAN_PERIOD | Periods of continuous health insurance coverage, including NHI used to characterize insurance status and payer type over time.<br>/ **Payer period start date, end date, payer concept (32725: NHI program) etc.** |
| COST | Reimbursed costs associated with visits, including NHI payment and patient out-of-pocket amounts. Derived from claim payment information.<br>/ **Cost domain (only 'Visit'), cost type concept (cost overall: 31985), currency concept (44818598: South Korean won), paid by payer, paid by patient etc.** |
| DRUG_ERA | Derived episodes of continuous drug exposure, created from DRUG_EXPOSURE records using standard OHDSI persistence rules, and used for long-term exposure and treatment pathway analyses.<br>**/ Drug concept, start date, end date, exposure count, gap days etc.** |
| CONDITION_ERA | Derived episodes of continuous disease, created by combining successive condition occurrences for the same condition concept within a pre-specified persistence window.<br>**/ Condition concept, start date, end date, occurrence count etc.** |

### Figure S1. Data Quality Dashboard result

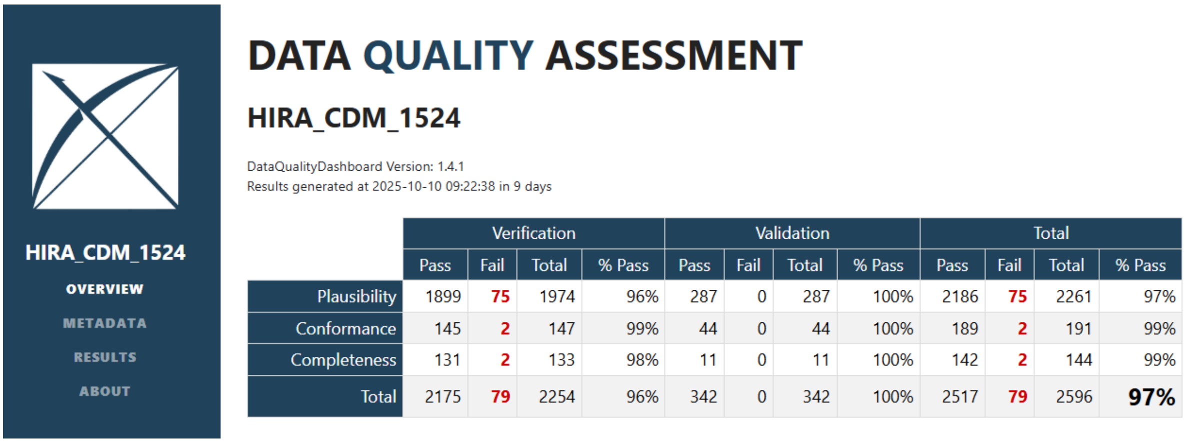


| | Verification | | | | Validation | | | | Total | | | |
|---|---|---|---|---|---|---|---|---|---|---|---|---|
| | Pass | Fail | Total | % Pass | Pass | Fail | Total | % Pass | Pass | Fail | Total | % Pass |
| Plausibility | 1899 | 75 | 1974 | 96% | 287 | 0 | 287 | 100% | 2186 | 75 | 2261 | 97% |
| Conformance | 145 | 2 | 147 | 99% | 44 | 0 | 44 | 100% | 189 | 2 | 191 | 99% |
| Completeness | 131 | 2 | 133 | 98% | 11 | 0 | 11 | 100% | 142 | 2 | 144 | 99% |
| Total | 2175 | 79 | 2254 | 96% | 342 | 0 | 342 | 100% | 2517 | 79 | 2596 | 97% |
